# My flight was cancelled! An exploratory study on institutional professional development budgets for medical educators

**DOI:** 10.1101/2025.09.15.25335775

**Authors:** Jeannine Nonaillada, Leigh Ann Holterman

## Abstract

**Introduction:** Recent federal funding cuts have created pauses in admissions, hiring, and execution of research studies at academic medical centers internationally. These monetary reductions have also impacted allowances for non-essential faculty travel. As a result, faculty may now be faced with challenges in how they obtain professional development.

**Methods:** A cross-sectional, exploratory study was implemented to discover the impact of funding cuts on faculty travel for professional development opportunities, as well as strategies medical educators are using to mitigate the current landscape.

**Results:** Findings indicate that faculty now have to use alternative methods to obtain professional development and that institutional guidance is lacking in how to do so.

**Discussion:** Authors provide concrete action steps for faculty to take amidst this challenge to remain engaged in professional development.

## Introduction

It has become the standard expectation for medical educators to contribute and produce scholarly works for career development and academic promotion [1]. Medical education conferences are one venue that hosts examples of these, including workshops, symposia, ‘quick shot’ podium presentations, and posters. Faculty and staff working in academic medical centers have traditionally attended medical education conferences for their own learning and professional development, as well as networking with national and international peers. These opportunities often require travel, with these costs covered by faculty development funds or institutional budgets.

Recently, however, funding cuts globally have led to drastic changes at academic medical centers. Specifically in the United States, pauses on admissions, hiring and research have been implemented [2 3]. The reach of these alterations has also extended internationally, with many countries in the world experiencing the loss of funding for biomedical research [4]. These funding cuts have affected monetary allowances to employees for what is deemed as non-essential travel [5 6]. As a result, faculty at academic medical centers globally may be faced with challenges in attaining professional development needed for growth and promotion, and thus risk losing engagement in this area.

### Methods

In this piece, we propose helpful action steps for faculty at academic medical centers to continue their own professional development amongst these current challenges.

To develop these action steps, we incorporated information from an exploratory study we conducted from June 1, 2025, until July 7, 2025. We designed an online survey to explore the current landscape of institutional professional development budgets for faculty in medical education and to learn strategies that were being used to mitigate changes in institutional professional development budgets. After institutional review board approval, we utilized a non-probability convenience sampling strategy by disseminating the survey to DR-ED, an electronic discussion group for medical educators, which is an established medical education listserv with over 4,000 subscribers, https://omerad.msu.edu/dr-ed-an-electronic-discussion-group-for-medical-educators. We also posted the survey invite to the virtual Community Pages of the Association of American Medical Colleges (AAMC) and International Association for Health Professions Education (AMEE), consisting of subjective and objective feedback. All data collected were anonymous, contained no personal health identifiers, and were analyzed in aggregate.

## Results

Our sample consisted of active subscribers to the virtual DR-ED discussion group and virtual members of the Group on Faculty Affairs (GFA) and Group on Educational Affairs (GEA) Community Pages of the AAMC, as well as the virtual members of the Faculty Development Special Interest Group of AMEE. We received 70 responses to our electronic survey. Of those that shared the name of their institution, none were from outside of the United States. It was hypothesized that recent federal funding cuts have impacted travel budgets for faculty professional development in academic medical centers.

Data were analyzed by descriptive statistics to obtain patterns and trends. Of those who responded to the survey item inquiring if their institution has a budget that enables attendance at professional meetings requiring travel, 74% (n = 49) answered ‘yes’. Further stemming from those same respondents, 71% (n = 35) stated these travel budgets have remained the same in the past 12 months, with only 29% (n = 14) reporting a decrease. From the respondents who reported either the absence of or a decreased budget for travel, when asked if their institution has offered any guidance on alternative methods for professional development, 94% (n = 29) responded ‘no’. However, these respondents shared many of their own alternative methods they are currently engaging in, and we also received numerous comments from survey respondents about the impact of their institution’s decrease in travel budgets for professional development meetings (Table 1).

**Table 1.** Major Themes in the Feedback on Mitigation and Impact of Decreased or Absent Travel Budget

| **Theme** | **Examples** |
| --- | --- |
| Virtual professional development | “Applied to virtual conferences.”<br>“I will focus on conferences I can attend online.” |
| Alternative institutional funding sources | “Apply for outside sources from Department including Provost Office.”<br>“I am working to cover some of the expense with expiring professional funds” |
| Self-funded travel | “I am funding travel out of pocket.”<br>“Once I start using personal funds, I'm afraid the college will make that something expected.” |
| Proactive networking | “Since much of the purpose of meetings is to connect with others in my field, I'm reaching out for 1:1 zoom meetings with specific people...”<br>“...I can still reach out to people I already know and maintain those relationships from a distance, but it's very difficult to develop new connections on-line” |
| Not submitting work to or attending conferences | “I am unable to attend, even a meeting at which I had 2 session proposals accepted.”<br>“Have not submitted any proposals.” |
| Promotion | “As educators, we are handicapped when we can't network at meetings and share our latest works.”<br>“It's ridiculous to work for an academic institution that requires national presentations for promotion and then not financially support that mission.” |

The majority of survey respondents (85%, n = 56) stated they plan to attend between 1-4 professional development meetings requiring travel in the next 12 months.

## Discussion

Survey findings support our hypothesis that recent federal funding cuts have impacted travel budgets for faculty professional development in academic medical centers. This has occurred in two ways: a decrease in institutional budgets for faculty travel to professional meetings, and a lack of formalized guidance from institutional leadership on either how to mitigate this change or find alternative methods for professional development.

The current landscape affecting institutional budgets for faculty travel to professional meetings, while it may be only temporary, has contributed to feelings among our survey respondents of this being indefinite, prompting faculty to consider the ways in which they obtain enrichment opportunities. For example, respondents shared they have to plan well ahead of the academic and fiscal year which meetings to attend so that they will be provided reimbursement. With limited budgets, faculty are choosing to opt out of extra meeting/event sessions that are not essential, such as receptions and pre-meeting gatherings that were previously attended. Some choose not to submit abstracts or attend conferences at all, which may have deleterious effects on career progression.

Respondents shared that virtual conferences and presentations are a useful option for professional development in the face of limited travel funding. However, faculty often rely on the networking and sharing of work among national peers that happens at in-person events, especially as these are often required for academic promotion and tenure [7 8]. While an aspect of these connections can be conducted virtually, it requires being proactive in order to achieve these interactions, when in contrast, they seem to occur organically when attending a meeting in person, especially in the form of spontaneous interactions. It is important to understand the benefits of virtual professional development while also recognizing what is lost when in-person events are no longer an option.

In an effort to maintain access to in-person conferences, some respondents reported they are using their own personal funds to cover travel and attendance at national meetings. However, we also received comments from respondents that they are hesitant to use their own personal funds as this could result in being the permanent expectation in academic medical centers for how faculty attend professional development meetings that require travel. This solution is inequitable, as many faculty may not possess the personal funding to cover these costs, especially junior faculty who may be most in need of these experiences.

Although we cannot be entirely certain, we postulate a contributing factor to our sample not naming any international institutions as their affiliation may speak to the large volume of public dissatisfaction with the substantial impact this is having currently in the United States as compared to other nations. We hope that this report will prompt other colleagues to continue exploring the impact of this issue internationally.

While acknowledging our study’s small sample size, we achieved saturation in our survey responses. Thus, the takeaway messages and major themes may be generalized to a wider audience of faculty who are affected by these cuts in federal funding and are now seeking alternative ways to obtain professional development and remain engaged.

We provide readers with the following action steps for faculty to take on seeking professional development opportunities:

1. Plan ahead before the start of the academic and fiscal year by acquiring the national and international academic meeting schedules. Compare costs to your faculty budget allowance to ensure that you prioritize conferences that will allow you to stick with your allotted budget set by administration.
2. Submit an abstract/workshop/poster to the meetings you would like to attend. Institutional administration may be more likely to cover travel costs if you are presenting at the meeting.
3. Take advantage of virtual meetings and webinars through organizations such as AAMC, IAMSE (International Association of Medical Science Educators), SACME (Society for Academic Continuing Medical Education), and AMA (American Medical Association), and AMEE to name a few. Although many of these are free, some do still require a registration fee albeit significantly less costly than a meeting with travel and lodging.
4. Join special interest groups/communities of practice within medical education organizations. Within these groups, scholarly collaborations may arise between institutions, which can be leveraged for opportunities for you to present at a Grand Rounds or to provide a workshop or lecture. These opportunities are impactful for your Educator Portfolio and promotion/tenure.
5. Apply for funding through grants (e.g., the AAMC GEA MESRE (Group on Educational Affairs Medical Education Scholarship Research and Evaluation), IAMSE Curriculum Innovation Grants, AMEE faculty development grants). Within your proposal, include funding for professional development/conference travel fees to disseminate your findings.
6. Make the most of professional development at your own institution. Many universities and hospital systems have free professional development courses and programs that faculty and staff can access. This might also include tuition remission for coursework.

## Conclusion

The recent changes to federal funding and the subsequent budget cuts at academic medical centers have clearly had an impact on the ability of faculty to travel for professional development. This has created

challenges in areas such as networking, knowledge growth, and faculty promotions. In turn, faculty are attempting to navigate what is potentially a new normal in the domain of professional development. However, the six recommendations above suggest alternative avenues to professional development opportunities that can help mitigate the negative effects of funding cuts and foster continued engagement among faculty. Until such a time that budgets return to their former status, we hope that these suggestions help our colleagues continue learning and growing in their careers.

### Takeaways

1. Faculty need to be proactive and plan professional development opportunities each academic year in accordance with institutional budget allowances.
2. Faculty should explore new opportunities for professional development to include virtual and local options that do not require travel.
3. Faculty must take advantage of networking opportunities and communities of practice to enhance venues for scholarly work to broaden professional development.

## Supporting information

IRB approval

## Data Availability

Raw data were generated at NYU Langone Health. Derived data supporting the findings of this study are available from the corresponding author [JN] on request.

## References

1. Cameron MW, Crowther LN, Huang GC. Faculty Development and Infrastructure to Support Educational Scholarship: A Scoping Review on Author Development. Acad Med 2023;98(1):112–22 doi: 10.1097/ACM.0000000000004896 [published Online First: 2022/08/04].

2. Davis CS, Wilkinson KH, Lin E, et al. Precision medicine in trauma: a transformational frontier in patient care, education, and research. Eur J Trauma Emerg Surg 2022;48(4):2607–12 doi: 10.1007/s00068-021-01817-7 [published Online First: 20211116].

3. Nietzel MT. More Universities Slow Spending, Admissions Over Federal Funding Chaos. Forbes, 2025.

4. Hudson RL. Trump halts new NIH grants to international health-research partners. Science Business: Science Business Publishing Ltd., 2025.

5. Schreiber M. Travel, grant and funding cuts ‘stifling’ US health agencies in new Trump era. The Guardian, 2025.

6. Mallapaty S. ‘All this is in crisis’: US universities curtail staff and spending as Trump cuts take hold. Nature, 2025.

7. Oh L, Linden JA, Zeidan A, et al. Overcoming barriers to promotion for women and underrepresented in medicine faculty in academic emergency medicine. JACEP Open 2021;2(6):e12552.doi: 10.1002/emp2.12552.

8. Rice DB, Raffoul H, Ioannidis JPA, Moher D. Academic criteria for promotion and tenure in biomedical sciences faculties: cross sectional analysis of international sample of universities. Bmj 2020;369:m2081 doi: 10.1136/bmj.m2081 [published Online First: 20200625].

