## Supplementary material for "My flight was cancelled! An exploratory study on institutional professional development budgets for medical educators": IRB approval

NYU Langone Health

Office of Science and Research
Institutional Review Board
1 Park Avenue | 6th Floor | New York, NY 10016

### Notification of IRB Review

May 29, 2025

On 5/29/2025 the IRB reviewed the following submission:

| | |
|---|---|
| principal investigator | Jeannine Nonaillada |
| study number | i25-00587 |
| study title | My flight was cancelled! Examining the current landscape of institutional professional development budgets for medical educators |
| Approval date | 5/29/2025 |
| location(s) | NY NASSAU - NYULH Hospitals |
| deparment | NYU Grossman Long Island School of Medicine |
| review type | Initial Study - Exempt 2(i) |
| board name | All boards |
| materials approved for use | • Exempt Protocol, Category: IRB Protocol;<br>• Key Information Sheet, Category: Consent Form<br>• Information Sheet, Category: Recruitment Materials;<br>• Survey, Category: Recruitment Materials;<br><br>Identifiable protected health information will not be used or disclosed in this research study therefore authorization to use protected health information is not applicable. |
| Link to Study | i25-00587 |

Thank you for using an NYU Langone Institutional Review Board (IRB). The current IRB Status of your submission is: Exempt.

This submission was reviewed the IRB and was determined to meet the criteria for exemption under 45 CFR 46.104(d).

Studies meeting the Exemption criteria do not require ongoing IRB oversight after the initial exempt determination. The Principal Investigator is responsible for ensuring this study is conducted in compliance with the materials submitted for this exempt determination and closing the study once all activities are complete. If there are changes that would impact the scope of this study or risks to participants or others involved in the research, please contact the IRB at 212-263-4110 or.

Your study cannot commence until all ancillary review decisions are complete. To determine the state of all ancillary reviews, go the MyStudies page of this study in Research Navigator. Ancillary review statuses are located on the top/right area of your study's main screen. Note: Ensure that approval has been issued in MyAgreements/CRMS and the Clinical Research Support Unit ("CRSU") before you proceed with any aspect of this study, including the enrollment of human subjects.

For questions regarding this submission, please contact David Decker (212-404-4100/).

29-MAY-2025

**Office of Science and Research**
**Institutional Review Board**
1 Park Avenue | 6th Floor | New York, NY 10016

29-MAY-2025

Thank you for using *NYU Langone Health IRBs*.

#### Review Notes

For NIH grant funded research approved before the revised Common Rule: the IRB has found the IRB approved protocol referenced above to be consistent with the NIH grant application.

NYU Grossman School of Medicine Federal wide Assurance: FWA00004952
NYU Winthrop/Long Island School of Medicine Federal wide Assurance: FWA00000726

*NYU Langone Health IRBs operate in accordance with Good Clinical Practices (GCP) and applicable laws and regulations. Federal rules allow IRBs to document their determination/authorization process in their policy manual. Determination letters generated by NYU Langone Health IRBs administration system are not physically signed as per policy. All approved study materials are clearly identified and locked in each study submission record within the IRB's administration system.*

#### *NYU Langone Health IRB Policy*

- *All current IRB policy documents can be found on our website*
- *You must submit all modifications to this study (e.g., protocol updates, modified recruitment materials, consent forms, etc.) using Research Navigator to communicate with the IRB ("eSubmission") for review and approval prior to initiation of those change(s), except where necessary to eliminate apparent immediate hazards to the subject(s). Changes made to eliminate apparent immediate hazards to subjects must be reported to the IRB within 24 hours.*
- *All adverse and/or unanticipated event(s) that occur while conducting this study must immediately be reported to the IRB via eSubmission.*
- *You may only use IRB-approved copies of your consent form(s), questionnaire(s), letter(s), advertisement(s), etc. in your study. Never use expired consent forms.*
- *If modifications are made to the study or adverse events occur while conducting study, the PI must inform all research staff listed on this study.*
- *IRB's approval is valid as per the period indicated above. A reminder to submit a continuation (should one be required) will be e-mailed to the PI, PI Proxy and Primary Contact 90, 60 and 30 days prior to this study's expiration date if one is indicated. After expiration, a daily reminder will be sent for 30 days followed by a weekly reminder until the study receives re-approval or a study closure.*
- *Prior to initiating an IRB-approved study, you must receive written approval from an authorized representative for each site where your study will take place. Key contacts are:*
    - Bellevue Hospital (BHC): if you are conducting all or part of your study at BHC, you must contact them to obtain additional approvals. BHC will be notified if any of their sites are selected as a location where your study takes place, but your team is obligated to contact them at to find out what approvals are required before conducting any research at a BHC location.
    - *CTSI - Clinical and Translational Science Institute, NYU School of Medicine [formerly General Clinical Research Center (GCRC)]:*
    - *NYU Langone Health Centers (Tisch Hospital/Rusk Institute/Co-op Care/HJD/Perlmutter Cancer Center) site approval is handled for you automatically (as needed) by the CRSU*
- *The IRB may suspend or terminate studies that are not in compliance with NYU Langone Health IRBs Policies & Procedures and the requirements of the Institution's Federal Wide Assurance with the federal government.*
- *Direct IRB questions and comments to 212-263-4110 or*

#### *Let Us Know How We're Doing*

*Click on the title above to send us feedback via a short, anonymous survey. Providing exceptional customer service is a top priority of the IRB and your responses will help us understand how we can continue to improve our service to the research community.*
